# Livestock-related deaths in Great Britain (2010-2023)

**DOI:** 10.1101/2025.02.27.25323016

**Authors:** Anna Chartres, Jo WH Oultram, Alex Royden, Sarah Shanks, John SP Tulloch

## Abstract

**Objectives:** This study aimed to describe the incidence, demographics, and context of livestock-related deaths 2010–2023.

**Study design:** A descriptive analysis of the Health and Safety Executive’s ‘Fatal injuries in agriculture, forestry and fishing in Great Britain’ annual reports.

**Methods:** Annual incidence of death was calculated. Demographic and contextual information were extracted, and data stratified by whether the victim was a farmer or member of the public. Statistical comparisons were made using Chi^2^ tests, Fisher’s exact test, and Mann Witney U Tests, where appropriate.

**Results:** Out of 78 livestock-related fatalities, 97% were caused by domestic cattle and 74.3% were farmers. Median age of all fatalities was 67 (range:29-87), and 63.4% were alone at the point of injury. Farmers were predominantly male and tended to be working with individual animals, often in a contained space. All members of the public were killed in fields and 84.2% had a dog present with them. They were 250 times more likely to have a dog present than farmers (OR=250, p<0.001). A calf or per-parturient cow was more likely to be present when a member of the public was killed (OR=4.3, p<0.01).

**Conclusions:** Cattle-related incidents are a concern and further research is required to enable the development and implementation of effective safety interventions. We recommend that a database of livestock-related injuries is created, measures are introduced so farmers can easily temporarily divert public rights of way, and educational programmes for farmers and the public are developed.

## Introduction

The ‘agriculture, forestry, and fishing’ sector has the worst rate of worker fatalities of any major industry sector in Great Britain (GB) (7.5 fatal injuries per 100,000 employees in 2023-24), averaging around 21 times higher than the all-industry rate.^1^ Within this sector, ‘injured by an animal’ is the second most common cause of death, after being ‘struck by a moving vehicle’.^2^ In the United States of America, cattle cause more occupational deaths than any other animal species.^3^ The GB Health and Safety executive (HSE), have stated that these fatal injuries are a huge concern to themselves and the agricultural industry.^2^ Despite this, there is little work exploring the demographics and context of those killed, or injured, by livestock.

Cattle, particularly bulls, are capable of causing damage to people with similar forces to those produced by motor vehicle collisions.^4^ In a UK study exploring emergency department attendance for cattle-related injuries, around 70% of patients required surgery, and 52% had an injury severity score (ISS) greater than 16.^5^ Other studies exploring hospitalised patients identified that 4-12% had ISS greater than 16.^6,7^ Trauma included concussions, facial fractures, closed head and chest trauma, and trauma at multiple locations.^6^ All studies showed a predominance of male patients with mean age in their late 40-50s. ^6–9^ One study highlighted that increasing age was associated with increased utilisation of care within hospitals. ^9^

Farmers work closely with cattle for a variety of practical and husbandry reasons. Moving and handling large animals in agricultural settings containing a substantial amount of concrete and metal (i.e. handling crushes, gates, pens) may increase the severity of injuries.^10,11^ Members of the public are also at risk of injury and death from cattle.^12^ These incidents tend to occur when walking, often with dogs, on public rights of way where cattle reside.

Current interventions to minimise risk from cattle-related injuries primarily focus on health and safety guidance,^13,14^ training of agricultural workers,^15,16^ and use of specialised equipment for handling cattle. Many farmers will rely on self-assessed capability, adapted by sociocultural norms and perceived consequences of certain behaviour, to adopt what they perceive are safe working practices.^17^ The countryside code provides guidance for the public around safe use of rural areas, including; signage, bans of certain bulls on public right of ways, and advice on keeping dogs under control.^18^

This study aims to describe demographic and contextual factors associated with livestock-related fatalities in Great Britain, and to identify any commonalities where injury prevention interventions may be targeted.

## Methods

Workplace fatalities, to either workers or members of the public, must be reported by law to HSE.^19^ This enables HSE to identify contextual information, trends in incidence, and to guide future policy and legislation, with the aim of preventing similar occurrences. The HSE publishes annual agriculture fatality records in the report, ‘Fatal injuries in agriculture, forestry and fishing in Great Britain’,^2^ in which a summary of each death provides some contextual information about the fatality.^20^

The HSE summary reports between the financial years 2010-2011 and 2022-2023 were compiled and information about livestock-related deaths were extracted; reports prior this were not available. Annual incidence of livestock-related deaths was calculated using two different denominator populations. Firstly, using the Office for National Statistic (ONS) population data as the denominator,^21^ and secondly by using livestock species specific population data.^22^ Incidence was stratified by each respective nation of GB.

The following demographic variables were extracted from each record: age, sex, and whether they were employed in agriculture or a member of the public. The following contextual information was extracted: animal species, sex of animal, whether the animal was in a group or isolated, the method of injury, location of incident, if the victim was alone, if the animals were being handled, if a dog was present, and whether a periparturient animal or infant animal was present. These variables were descriptively analysed. Subsequently data were stratified based on whether the victim was employed in agriculture or a member of the public. Statistical comparisons between these two groups were made using Chi^2^ tests, Fisher’s exact test, and Mann-Witney U Tests, dependent upon the variable’s characteristics and data structure.

All statistical and spatial analyses were carried out using R language (version 3.2.0) (R Core Team 2015). Results were deemed statistically significant where p < 0.05. No ethical approval was needed as this was an analysis of publicly available data, with no personally identifiable information.

## Results

Between 2010 and 2023 there were 78 livestock-related fatal injuries in Great Britain. All involved bovids, 97.4% (95% CI 91.0-99.7, n=76) of fatalities involved domestic cattle, whilst the other two deaths involved water buffalo. Due to the predominance of domestic cattle in the fatalities, the remainder of the analysis will focus solely on this species.

There was no overall trend in incidence in GB during the study period. The mean annual incidence was 0.9 deaths per 10 million population, this represents six deaths annually, ranging from 0.3 to 1.4 deaths per 10 million population (Fig 1). The lack of trend was similar across the three nations, the mean annual incidence in England was 0.8 deaths per 10 million population, in Scotland 1.3 deaths per 10 million population, and in Wales 1.5 deaths per 10 million population.

**Figure 1.**
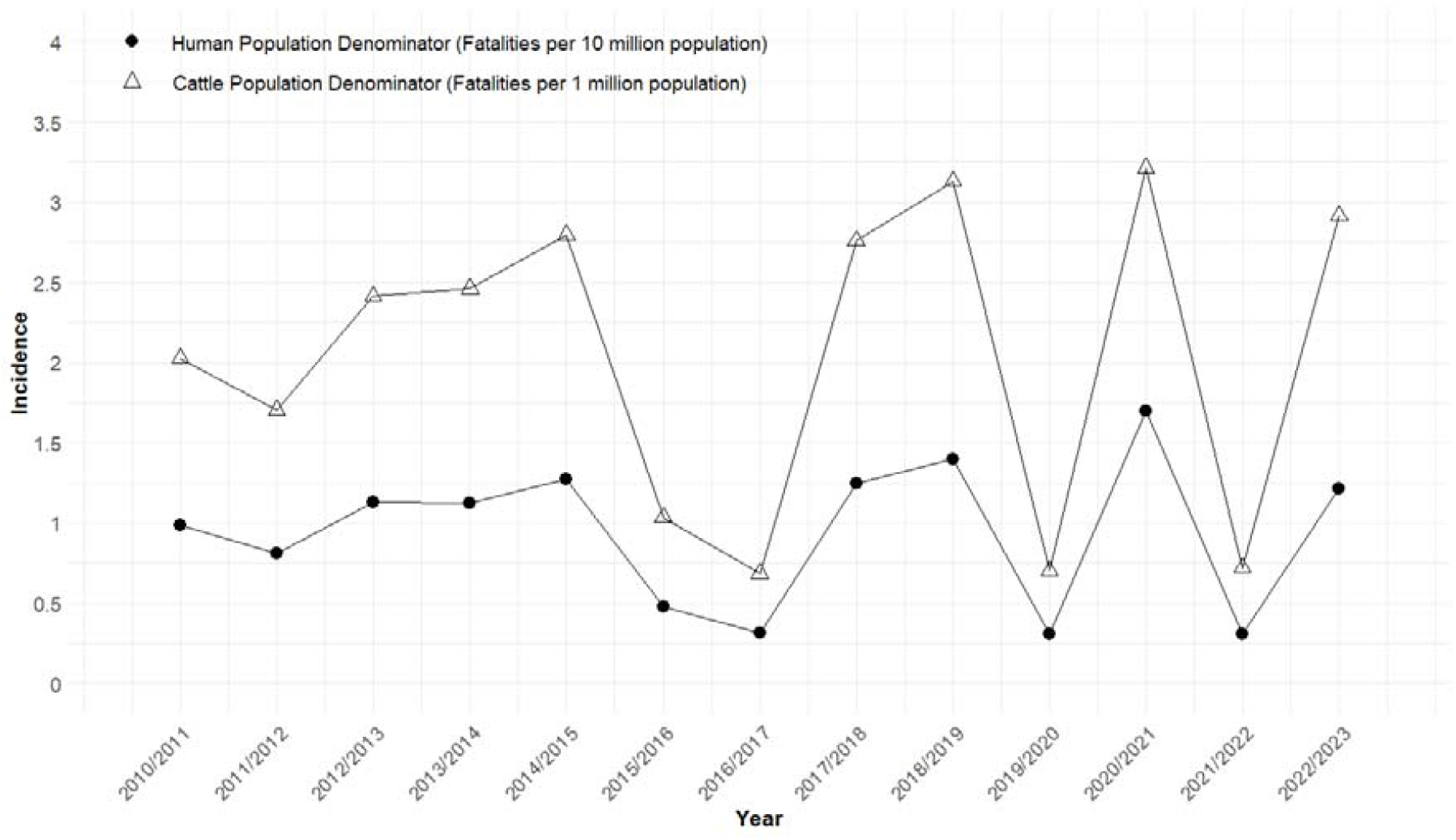
Incidence of cattle-related fatalities, with humans and cattle as the denominator population, in Great Britain (2010-2011 and 2022-2023)

Similarly, there was no overall trend in incidence in GB when cattle population was used as a denominator, the mean annual incidence was 2.0 deaths per 1 million cattle population. The lack of trend was similar across the three nations, the mean annual incidence in England was 2.5 deaths per 1 million cattle population, in Scotland 1.2 deaths per 10 million population, and in Wales 1.1 deaths per 10 million population. For context, in 2023 there were 1.7 million adult cows in England, 0.6 million in Scotland, and 0.4 million in Wales.^22^

### Demographic and contextual information

Two records from 2016/2017 had no summaries attached and were therefore excluded from further analysis, leaving 74 fatality records for analysis. Of these, 74.3% (95% CI 62.8-83.8) were farmers (n=53), agricultural contractors (n=3) or farming family members (n=1). These have been deemed ‘farmers’ for the remainder of the analysis. The remaining 25.7% (95%CI 16.2-37.2) were members of the public. The relative proportion of fatalities between the two groups does not appear to have changed over time (Fig 2).

**Figure 2.**
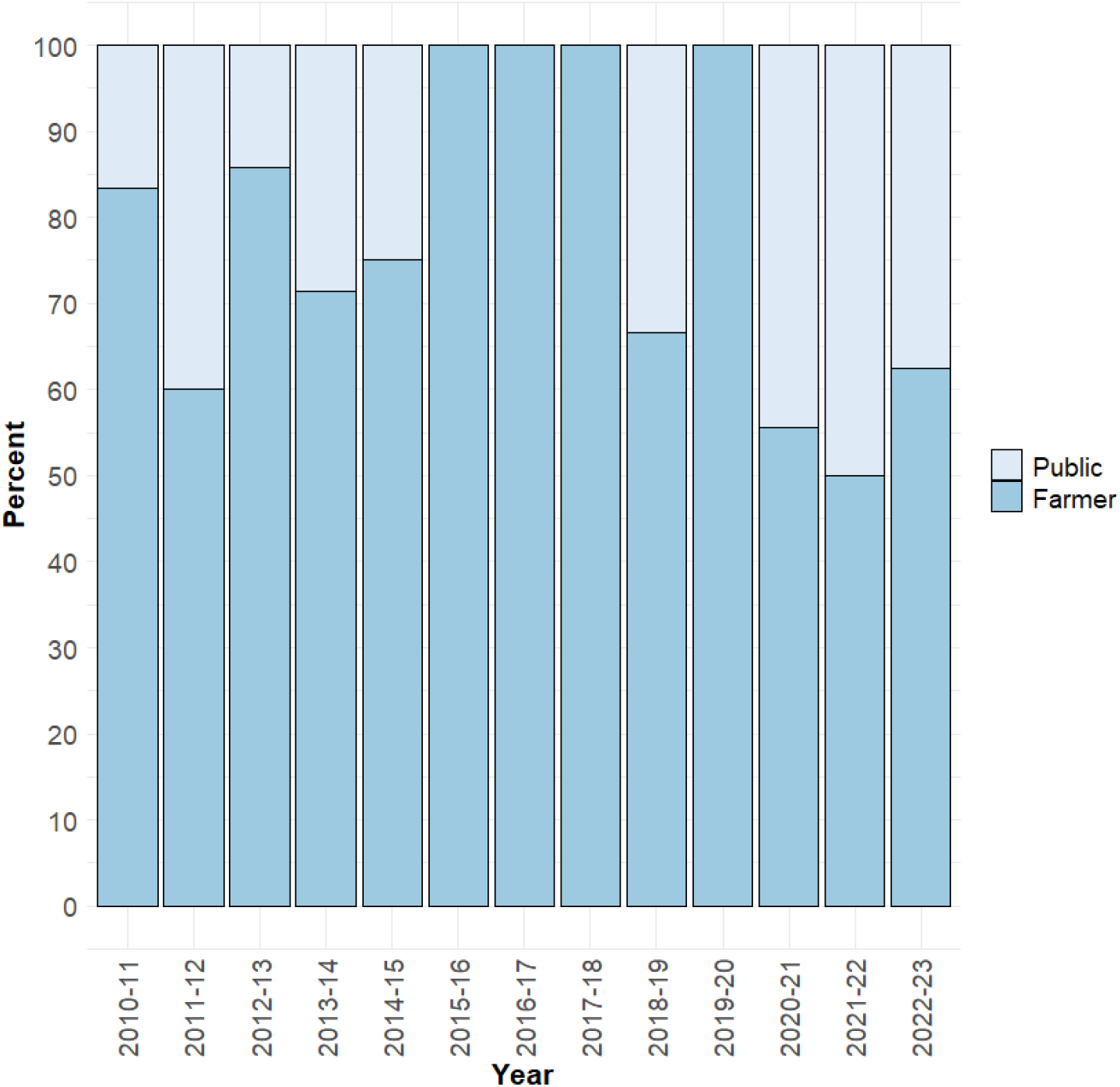
Relative proportion of farmers and the public killed by cattle in Great Britain (2010-2011 and 2022-2023)

Victims were predominately male, and farmers were seven times more likely than members of the public to be male (Table 1).

**TABLE 1.** Demographic and contextual variable of farmers and the public killed by cattle in Great Britain (2010-2011 and 2022-2023) [N/C = Not calculable, N/A = Not applicable, OR=Odds Ratio].

| Variable | Overall | Farmers (n=55) | Public (n=19) | Statistical Test | Estimates | Significance |
| --- | --- | --- | --- | --- | --- | --- |
| **Sex (% Male)** | 82.4% (71.8-90.3) | 90.9% (80.1-97.0) | 57.9% (33.5-79.8) | Chi<sup>2</sup> test | OR=7.0<br>(95%CI 1.9-27.9) | p<0.01 |
| **Age (median and range)** | 67 (29-87) | 67 (33-87) | 66 (29-85) | Mann-Witney U Test | z-score = 0.39 | p=0.70 |
| **Animals involved.**<br>**(as defined by the Health and Safety Executive)** |  |  |  | N/C |  |  |
| Bull | 21.6% (12.9-32.7) | 29.1% (17.6-42.9) |  |  |  |  |
| Bulls | 1.4% (0.03-7.3) | 1.8% (0.05-9.7) |  |  |  |  |
| Bullocks | 2.7% (0.3-9.4) | 3.6% (0.4-12.5) |  |  |  |  |
| Cow | 28.4% (18.5-40.1) | 38.2% (25.4-52.3) |  |  |  |  |
| Cattle/Cows | 32.4% (22.0-44.3) | 21.8% (11.8-35.0) | 63.2% (38.4-83.7) |  |  |  |
| Cattle + Bull | 13.5% (6.7-23.5) | 5.5% (1.1-15.1) | 36.8% (16.3-61.6) |  |  |  |
| **Calf or peri-parturient cow present** | 41.9% (31.3-53.3) | 32.7% (21.8-46.0) | 68.4% (45.8-84.8) | Chi<sup>2</sup> test | OR=0.23<br>(95%CI 0.07-0.70) | p<0.01 |
| **Location (n=69)** |  | n=50 | n=19 | N/C |  |  |
| Contained space | 46.3% (34.3-58.8) | 64.0% (49.2-77.1) | 0% |  |  |  |
| Field | 53.6% (41.2-65.7) | 36.0% (22.9-50.8) | 100% |  |  |  |
| **Alone at time of death (n=41)** |  | n=26 | n=15 |  |  |  |
| Yes | 63.4% (48.1-76.5) | 61.5% (42.5-77.6) | 66.7% (41.5-85.0) | Chi<sup>2</sup> test | OR=0.8<br>(95%CI 0.2-3.1) | p=0.76 |
| **Dogs Present** | 23.0% (14.8-33.8) | 1.8% (0.0-10.5) | 84.2% (61.6-95.3) | Fisher's exact test | OR=0.004<br>(95%CI 0.00008-0.04) | p<0.0001 |
| Moving, handling, or treating cattle (n=52) |  | 67.3% (52.9-79.7) | N/A | N/A |  |  |

The median age was 67 and did not differ significantly between farmers and the public. Over a third of cattle fatalities (39.2%) were associated with male cattle (bulls or castrated bullocks). Farmers were predominately dealing with a single bull (29.1%), whilst the public were killed when a bull was in a field with cows (36.8%). More farmer fatalities occurred with solitary animals (67.3%) than groups. However, all public fatalities were associated with a group of animals. Over 40% of fatalities occurred either when a calf was present (n=26) or in the presence of a peri-parturient cow (n=5). Members of the public were 4.35 times more likely to have been killed in the presence of a calf or peri-parturient cow than farmers. It was only described in three incidents whether the cattle were a beef or dairy breed (2 dairy bulls and one beef cow). Farmers were predominately killed in a contained space (such as a yard, milking parlour, bull pen) (64.0%), whilst all members of the public were killed in fields. Of the records (n=42) that stated if the victim was alone or not at the time of the accident, 63.4% were alone. Sixty-seven percent of farmers were handling cattle at the time of the incident. Eighty-six percent of these incidents (85.7%, n=30) had enough information to identify the events preceding the injury. Forty-three percent (43.3%) were herding cattle, 20% were handling cattle in a crush or race, 20% were moving cattle on or off a trailer, and 16.7% were administering drugs to a cow (60% of these cows had a calf present). Eighty-four percent of public fatalities occurred when a dog was present and were 250 times more likely to have a dog present compared to farmer fatalities.

The most prevalent injury was trampling (50.0%), which was more prevalent in the public (73.7%) than in farmers (41.8%). Of note is that 18.8% (n=10) of farmers died of head injuries resulting from hitting concrete floors or walls, rather than direct injury caused by the cattle. However, method of injury was not specific and many of the descriptors (attack, butting, crushing, dragging, goring, indirect contact, kick, and knock to the ground) were used interchangeably.

## Discussion

Fatalities caused by livestock are a constant rare event in Great Britain, almost all related to domestic cattle, with the majority occurring to farmers. The incidence of livestock-related deaths is 1.5 times higher than that of dog-related deaths, ^23^ yet they do not receive the same media attention, related discussions, or push for legislative change. Farming victims tend to be older males, who are moving/handling solitary animals by themselves in a confined space (i.e. yard, pen). Public victims are elderly individuals, of either sex, who are solitarily walking their dog through a field containing a group of cattle, typically with calves, periparturient cow, or bull present.

Agricultural victims were predominately male, and this is likely a reflection of the sector, being 84% male.^24^ There may also be gender differences in cattle exposure as women are known to work fewer hours on farm,^25,26^ inhabiting traditional gender roles and are less likely to be working with cattle,^27^ have been shown to be more safety conscious, and more risk averse.^27,28^ The median age was also reflective of farmer demographics where 68% are over the age of 55.^24^ There are many common age-related conditions and syndromes such as cataracts, hearing loss, osteoarthritis, and frailty which may cause elderly farmers to experience increased risk of fatal injury and reduced ability to escape when the risk is posed.^29^ Additionally, the high degree of trauma experienced in cattle-related injuries means that increasing age decreases survivability.^8^ A UK study found that 21% of farmers plan to never give up working, putting them in potentially dangerous situations as they age and become vulnerable to injury.^30^ Injury prevention methods should be targeted at older, male farmers and there should be encouragement of retirement plans for older farmers to reduce the risk of livestock related fatalities.

This work highlights certain farm management practices may increase the risk of trauma, such as herding, working with crushes, races, and trailers, and performing procedures on calves when a mother is present. Almost 20% of farmers died of head injuries resulting from hitting concrete following contact with cattle, rather than from direct injuries sustained from the cattle. Agricultural environments require that building materials are strong, durable, and easily cleaned, yet this poses a hazard to the worker if they impact them with force. Exploring redesigning materials, equipment or farm layout may be needed to reduce risk. Furthermore, the adoption of wearing protective headgear could be explored.

Even experienced farmers must be vigilant with health and safety measures when around cattle and avoid risk taking as cattle behaviour can be unpredictable. Many cases in this study demonstrated inappropriate handling methods and dangerous actions. Additionally, these data suggest that many farmers were working alone at the point of their death. It is unknown whether their injuries would have been survivable if someone had been present to call emergency services; highlighting the dangers of lone-working in a high-risk profession.^31,32^

Deaths of farmers have a profound effect on others, as well as their business. A study in 2016 discovered that 55% of farm workers were family members and that 30% of the family workforce worked on the smallest farms, and only 14% on the largest farms.^33^ Families affected by a sudden farming death have been shown to take a pragmatic approach, and continue their farming activities, whilst accepting, and normalising, the risks inherent in farming.^34^ This potentially minimises their approach to health and safety on the farm and could lead to a ‘this is how we’ve always done it’ approach.

Contrasting to farmers, males were no more at risk than females whereas the age of those killed was similar to farmers, this possibly being linked to the same age-related conditions predisposing to injury/death. The average age of rural inhabitants is higher than those in urban areas, thus older people are more likely to live in areas where cattle are present.^35^ They also spend more time outside during the working week and summer than younger adults, again more like to encounter more cattle.^36^ Many of the individuals were by themselves when injured, this means that a significant amount of time could have elapsed before they were found or before emergency medical services were on the scene. Survivability of blunt and penetrating trauma is increased if emergency services can be on the scene within the hour and ideally transported to hospital by helicopter.^37,38^ This highlights the need for walkers to inform others of their itinerary when walking alone in the countryside.

All public fatalities occurred in fields, likely due to their access to public footpaths containing cattle and lack of public access to farm buildings. Similarly, there were no incidents that involved a single animal. The public were significantly more likely to have been killed in the presence of a calf or peri-parturient cow than farmers. Cows’ innate protective responses when calves are present, or around parturition, and if they feel threatened can result in aggression behaviour.^39^ This protective response is heightened when dogs are present. In over a third of cases a bull was present in a group of cows. Certain breeds of bulls are banned from being kept in fields with a public right of way.^40^ It is unclear why specific breeds have been singled out, as the HSE state that, ‘There remains no clear picture pointing towards any breed or breeds being more hazardous’.^41^

These data show a significant presence of dogs in deaths of the public (84.2%), which is supported by a previous study on dog involvement in cattle attacks (non-fatal and fatal).^12^ The data did not show what role the dogs played in the fatality (e.g. whether they attacked the cattle, whether the dog was loose, or whether it was under control) nor the owners subsequent behaviour. This information is critical to gain a greater understanding of the role that dogs play in fatal human-cattle interactions.

The main limitation of this study is its reliance on summary data analysis. According to HSE, the data contained in the reports had been “significantly simplified and therefore may not accurately describe the full circumstances”.^2^ This is problematic, as these are the only publicly available data describing these incidents. The data is inconsistently recorded, with limited contextual information. For example, they do not differentiate between dairy or beef cattle or cattle breed, animal groupings were unclear, cattle terminology relating to age and sex were used inconsistently, and descriptions around events preceding the fatality were often scant.

We have three recommendations, in addition to the advice provided by the HSE,^13,16^ and the countryside code.^14,18^ Firstly, to build a database of all livestock-related injuries nationally. There is currently no system for reporting non-fatal cattle-related injuries and as such data is restricted to what is reported to the HSE. A database would provide more contextual information about events preceding an injury or fatality and could aid in the development of injury prevention strategies.^42^ The addition of livestock-related injuries to medical coding systems such as ICD-10 or SNOMED codes should be explored. This would enable surveillance of cases which have been admitted into hospital and give a better understanding of the scale of the problem and the cost and burden to society. These codes are present for dog bites, and have done a lot to help raise the awareness of the scale of that issue.^43^ Without an understanding of the incidence of livestock-related injuries it will always be hard to develop, and justify, risk mitigation strategy developments. Further research into the context of livestock-related injuries is urgently needed.

Secondly, farmers should be able to temporarily divert the public rights of way around or away from a cattle field if they show that their livestock could not be feasibly grazed elsewhere. Farmers are currently encouraged to provide alternate routes, but the public are still entitled to use the right of way and so the risk remains.^13^ Temporary diversions are coordinated via a local authority following proper legal procedure.^40,44^ This process involves public consultation, is costly, and may take months to be approved. Furthermore, it must be repeated each time a temporary diversion is needed. Legislative changes or the introduction of measures that would allow more rapid installation of temporary diversions of public rights of ways through fields containing potentially dangerous livestock and aid public safety are required. This should be flexible and repeatable so that farmers can better reduce the likelihood of interactions between their livestock and the public.

Finally, further education is needed for both farmers and public. This must not be seen as the sole safety intervention but a key part of a suite of interventions. It needs to be applied consistently and repeatedly to have significant impact. Interestingly, educational interventions where older farming family members taught a set curriculum to younger family members were shown to have a positive impact in terms of knowledge and behaviours on both generations.^45^ With no single co-ordinating body, educating the public about safety around livestock will require a multi-organisational approach (i.e. HSE, Natural England, National Farmers Union, Rambler Association) with an array of outreach modalities. Joint farmer-public safety programmes could be a potential avenue for development. Particular focus needs to be paid to dog walkers, as so many public fatalities had a dog present; this could be facilitated by the dog charity sector.

## Conclusion

The number of fatal injuries within the agricultural sector is alarming and warrants further research. Whilst these deaths have huge impacts on individuals, preventative change has been slow. The data here likely represent the tip of the iceberg, as to date no research has explored national levels of hospitalisation or long-term impact of livestock-related injuries to individuals, families, or businesses. Systematic reporting of livestock injuries and research of circumstances leading up to injury is urgently needed for the design and implementation of better safety interventions, and to protect people from fatal livestock-related fatal injuries.

## Data Availability

All data produced in the present study are available upon reasonable request to the authors

## Author Statements

### Funding

This research did not receive any specific grant from funding agencies in the public, commercial, or not-for-profit sectors.

### Ethical Approval

No ethical approval was needed, as this was an analysis of publicly available data, with no personally identifiable information. Patients were not involved in the design, nor participated in the research, of this study.

### Declaration of interests

All authors have no conflicts of interest to declare.

### Author contributions

J.T. conceived and designed the work. AC and JT analysed the data and drafted the article. All authors interpreted the data, helped to revise the article and approved the final submitted article. All authors agree to be accountable for the accuracy and integrity of this work.

## Notes

### Competing Interest Statement

The authors have declared no competing interest.

### Funding Statement

This study did not receive any funding

